# Reinforcement learning-based control of epidemics on networks of communities and correctional facilities

**DOI:** 10.1101/2025.05.10.25327262

**Authors:** Christopher Weyant, Serin Lee, Jeremy D. Goldhaber-Fiebert

## Abstract

**Background:** Correctional facilities can act as amplifiers of infectious disease outbreaks. Small community outbreaks can cause larger prison outbreaks, which can in turn exacerbate the community outbreaks. However, strategies for epidemic control in communities and correctional facilities are generally not closely coordinated. We sought to evaluate different strategies for coordinated control and examine their robustness.

**Methods:** We developed a stochastic simulation model of an epidemic spreading across a network of communities and correctional facilities. We parameterized it for the initial phases of the COVID-19 epidemic for 1) California communities and prisons based on community data from covidestim, prison data from the California Department of Corrections and Rehabilitation, and mobility data from SafeGraph; and 2) a small, illustrative network of communities and prisons. For each community or prison, control measures were defined by the intensity of two activities: a) screening to detect and isolate cases and b) non-pharmaceutical interventions (e.g., masking and social distancing) to reduce transmission. We compared the performance of different control strategies including heuristic and reinforcement learning (RL) strategies using a reward function, which accounted for both the benefit of averted infections and non-linear cost of the control measures. Finally, we performed analyses to interpret the optimal strategy and examine its robustness.

**Results:** The RL control strategy robustly outperformed other strategies including heuristic approaches like those that were largely used during the COVID-19 epidemic. The RL strategy prioritized different characteristics of communities versus prisons when allocating control resources, and exhibited geo-temporal patterns consistent with mitigating prison amplification dynamics.

**Conclusion:** RL is a promising method for controlling epidemic spread on networks of communities and correctional facilities, providing insights that can help guide policy.

**Highlights:**

- For modelers, we developed a stochastic simulation model of an epidemic spreading across a network of communities and correctional facilities, and we parameterized it for the initial phases of the COVID-19 epidemic for California communities and prisons in addition to an illustrative network.
- We compared different control strategies, and we found that reinforcement leaning robustly outperformed the other strategies including heuristic approaches like those that were largely used during the COVID-19 epidemic.
- For policy makers, our work suggests that they should consider investing in the further development of such methods and using them for future epidemics.
- We offer qualitative insights into different factors that might inform resource allocation to communities versus prisons during future epidemics.

## Background

Correctional facilities, such as prisons and jails, have historically had substantially higher incidence rates of respiratory infectious diseases as compared to surrounding communities. For example, the COVID-19 incidence rate ratio for prisoners versus the general population in the United States (US) was estimated to be over 5 during the early phases of the pandemic.^1,2^ COVID-19 incidence rates were similarly very high in US jails and Immigration and Customs Enforcement (ICE) facilities.^3,4^ These trends hold for other respiratory infectious diseases, such as tuberculosis (TB) and influenza, and other countries, such as those in South America.^5,6^

Due to these higher rates, correctional facilities can act as amplifiers of respiratory infectious diseases. For example, Reinhart and Chen identified jail-community cycling as a significant predictor of community cases at the ZIP code level by analyzing COVID-19 cases in Cook County Jail and the general population in Chicago.^7,8^ The authors also identified a negative association between jail decarceration and community cases at the US county level.^9^ Malloy *et al.* found county-level measures of COVID-19 were a promising predictor of COVID-19 outbreaks in correctional facilities by analyzing data from 24 prison facilities in the Pennsylvania Department of Corrections and US counties.^10^ For a range of respiratory infectious diseases including variants of COVID-19, Weyant *et al*. used simulation modeling to illustrate a magnification-reflection dynamic. Small community outbreaks can cause larger outbreaks in correctional facilities, which can in turn exacerbate the community outbreaks.^11^ The authors also identified key factors governing the size and timing of this dynamic.

Despite this interdependence, epidemic control efforts in communities and correctional facilities are generally not closely coordinated. For example, during the COVID-19 epidemic, communities and correctional facilities generally implemented control measures primarily based on leading-edge measures of incidence within their own jurisdictions or else followed state-wide policies. This paper focuses on several key questions. First, for a network of communities and prisons, how do different control strategies differ in performance in terms of averting infections and costs? Second, does the optimal identified policy offer any qualitative insights into how control measures should be implemented and adapted more generally? Third, how robust is the optimal strategy to deviations from following the recommended actions exactly and to incorrect initial estimates of disease parameters?

To address these questions, we developed a simulation model of an epidemic spreading across a network of communities and correction facilities. Using both a network of California communities and prisons as well as a small, illustrative network of communities and prisons, we compared the performance and robustness of different control strategies including heuristic strategies and RL strategies. Finally, we examined the optimal strategy to derive qualitative insights.

## Methods

We developed a stochastic, metapopulation model of an epidemic on a network of communities and correctional facilities. We parameterized the model for the initial phases of the COVID-19 epidemic for 1) California counties and prisons and 2) an illustrative network of communities and prisons (Figure 1). For both networks, for control strategies including heuristic and RL strategies, we projected outcomes including infections and cost of the control measures for the first year of the epidemic. We also performed analyses to interpret the optimal strategy and investigate its robustness. Simulation of the epidemic and RL were conducted using Python, while analysis of simulation output and plotting were conducted using R. Input data and code for replication and extension of our analysis will be available via GitHub concurrent with publication.

**Figure 1.**
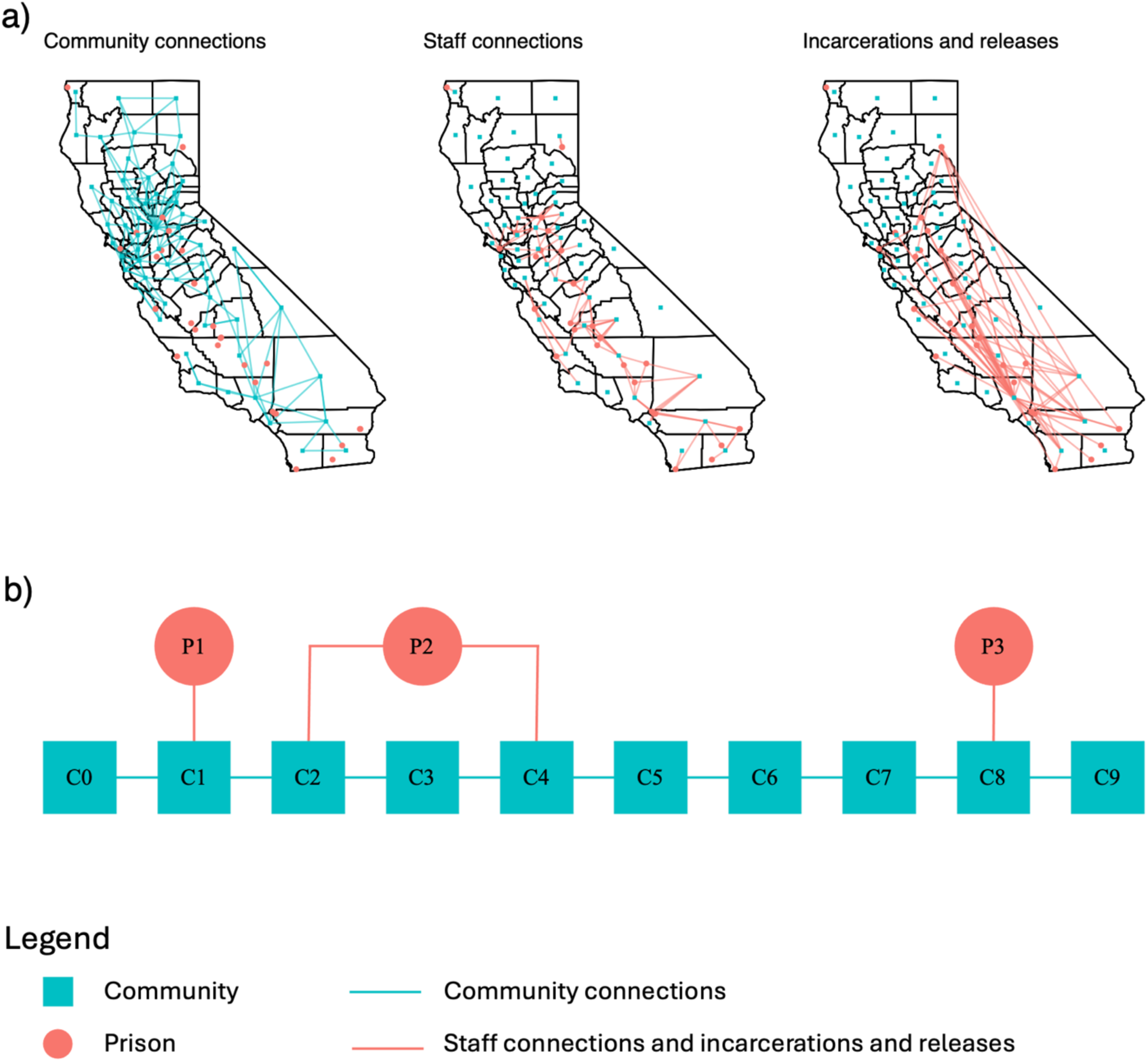
Network structures. Panel a shows the network structure for the California network while panel b shows the network structure for the illustrative network. The blue squares denote communities while the red circles denote prisons. For the California network, we only show the strongest connections to aid with figure readability. For the illustrative network, lines between communities denote community connections, while lines between communities and prisons denote connection by both staff and incarcerations and releases. Underlying these networks, we model epidemic spread using stochastic SEIDR models (Figure S1) for incarcerated people in each prison, correctional staff by the prison in which they work and their home community, and free-living people in each community. These models are not isolated, and people interact with each other based on the network connections as described in Text S1.

### Simulation model

We developed a stochastic, metapopulation susceptible-exposed-infectious-detected-recovered (SEIDR) model of infection transmission occurring within and between a set of communities and correctional facilities. We modeled SEIDR health states over time for incarcerated people in each prison, correctional staff by the prison in which they work and their home community, and free-living people in each community. We modeled movement of people between the communities each day (e.g., commutes) in addition to incarcerations and releases to/from prisons. Each community or correctional facility could implement two control measures—testing and non-pharmaceutical interventions (NPIs)—at six possible intensity levels (0%, 20%, 40%, 60%, 80%, and 100%). We simulated infections using the tau-leap method, which is an approximate method for stochastic simulation based on Gillespie’s algorithm, with a time step of a tenth of a day.^12^ Model outcomes included infections and costs based on use of the control measures. See Text S1 and Figure S1 for more detailed information regarding the simulation model.

### Model parameterization

We parameterized the simulation model for two cases, which served different purposes. First, we parameterized the model for the network of California counties and prisons during the early phases of the COVID-19 epidemic. This case was intended to yield insights on the relative performance and behavior of different control strategies in a context similar to ones in which they might be deployed. We determined populations of incarcerated people (by prison) and staff (by prison and home county) based on data from CDCR.^13^ We determined populations of people in each community based on data from the US census.^14^ We determined epidemiologic parameters based on published literature (Text S2, Table S1). We determined the mobility of free-living people from their home community to the other communities based on data from SafeGraph (Text S3), which tracks movement of millions of people with mobile devices in the US.^15,16^ We set illustrative costs for the control measures and a willingness-to-pay (WTP) threshold for infections averted (Text S4, Table S2). The costs were non-linear to reflect that it becomes ever more costly to achieve higher intensities of control measures as additional supplies are needed and/or people less likely to comply need to be convinced. We selected illustrative values as we seek to evaluate different control strategies, and the appropriate cost of an infection is a policy decision beyond the scope of this paper. We picked the values we did in part because they avoided the trivial solution of the optimal strategy being to use full mitigation or no mitigation. At the start of the model, we seeded 100 infections in Los Angeles County to simulate the start of an outbreak at a location that has high density of people and travelers; of note, our method can be used with any current distribution of infections.

Second, we parameterized the model for an illustrative example of a small number of communities and prisons. This case was intended to provide additional credibility to our findings from the California network and to allow for easier interpretation of the strategies. In this case, all nodes of a certain type (i.e., prison or community) were the same other than their connections, which again allowed for easier interpretation. Other parameters were generally the same as for the California network (Table S1).

### Heuristic strategy

Each community or prison implemented control measures at a certain intensity whenever the estimated per-capita incidence of infections in that community or prison was greater than a community or prison-specific threshold. There was one threshold and one intensity for all communities, and there was one threshold and one intensity for all prisons. To estimate the per-capita incidence of infections (detected and undetected), we adjusted the per-capita incidence of cases (i.e., detected infections) using the testing rate (Text S5). We did not directly compare the per-capita incidence of cases to a threshold as more testing would lead to more cases. We did not directly use the true per-capita incidence rate recorded during the simulation as this would be unknown during a real-life epidemic. We assumed some free, low-level baseline rate of detection for this strategy and all other strategies including RL to avoid the case where the no testing suggests no infections. We determined the thresholds and control measure intensities for communities and prisons, respectively, through exhaustive search (Text S6). As it takes time to collect data, interpret data, and promulgate policy changes, we assumed use of the control measures could only be updated once a week. We made this same assumption for the RL strategy.

### Reinforcement learning strategy

We implemented our environment using Gymnasium.^17^ Each episode was one year and consisted of fifty-two, one-week steps. The action space was multi-discrete, and, for each community or prison, allowed for two actions (one for testing and one for NPIs) each at one of the six possible intensity levels. The state space was continuous and tracked information for each community and prison on the per-capita incidence of cases (i.e., detected infections), the actions from the last step (necessary, for example, as interpretation of the incidence of cases depends on the testing rates), and estimated per-capita cumulative infections thus far in the epidemic. See Text S5 for how we estimated cumulative infections based on the incidence of cases and the testing rate. The reward was equal to the negative of the sum of the true number of infections and cost of the control measures converted to units of infections using a WTP threshold per infection. We used the true number of infections in the reward function but not the state space as we wanted the agent to recommend actions as intelligently as possible given the available information.

We used proximal policy optimization (PPO) and Stable Baselines3 (SB3) to learn policies.^18,19^ Hyperparameter tuning was conducted with Optuna and details can be found in Text S7 and Table S3.^20^ Learning was conducted for 3 million steps, which was sufficient for the reward to effectively stop improving in all cases.

### Analyses

We projected outcomes over the first year of the epidemic 1,000 times for each of four control strategies for both networks. These strategies included all communities and prisons implementing both testing and NPI control measures at maximum intensity for the entire model duration, no communities or prisons implementing control measures for the model duration, the heuristic policy, and the RL policy. We converted both infections and control measure costs to units of infections using a WTP threshold and summed them. The best strategy was the one that minimized the expected value of this cost function over the 1,000 replications.

To interpret the best policy for the California network, we calculated the average expenditure on control measures per person per week per control measure for each community and prison. We then fit XGBoost models using the caret package in R to predict the average expenditure for communities and prisons, respectively, as a function of their characteristics such as beta, size, and degree of connectedness.^21^ We selected characteristics such that they had low correlation with each other. From these models, we generated variable importance and partial dependence plots. These plots provided information on the relative importance of the variables in predicting outcomes and their marginal effects on the outcomes. We used XGBoost models as they gave better performance than other predictive models such as glmnet and random forest models (Table S4, Table S5).^22,23^

To interpret the best strategy for the illustrative network, we adopted a different approach. As there was no heterogeneity in the community and prison variables and as the dynamics of the illustrative network were not prohibitively large and complex, we relied on interpretations of visual trends instead of using XGBoost models as we did for the California network. To do so, we visualized epidemic curves and corresponding actions over time for each community and prison.

We determined the robustness of the control strategies by repeating the projection of outcomes under two alternative scenarios. In the first, to simulate non-perfect implementation, we varied each action that the control strategy recommended such that there was an equal chance of using one level less of mitigation (e.g., −20%), the recommended level, and one level more of mitigation (e.g., +20%). We enforced bounds such that the level could never be below 0% or above 100%. If the varied level would be less than 0% or greater than 100%, it was set to 0% or 100%, respectively. In the second, to simulate potential misestimation of the epidemic parameters early in the epidemic, we parameterized gamma distributions for the disease parameters such that two standard deviations was equal to 20% of the initial parameter value. We sampled parameter values from all these distributions at the start of each of the 1,000 replications.

The funding sources had no role in the study.

## Results

### California network

The RL strategy offered substantially better performance compared to the other control strategies for the California network (Figure 2a). We projected that following control all, control none, heuristic, and RL strategies would result in costs of 39.43, 30.96, 18.34, and 12.07 million infections, respectively. These costs are the sum of the actual infections and the financial costs of the control measures, which were first converted to units of infections using a WTP threshold as described in the Methods section.

**Figure 2.**
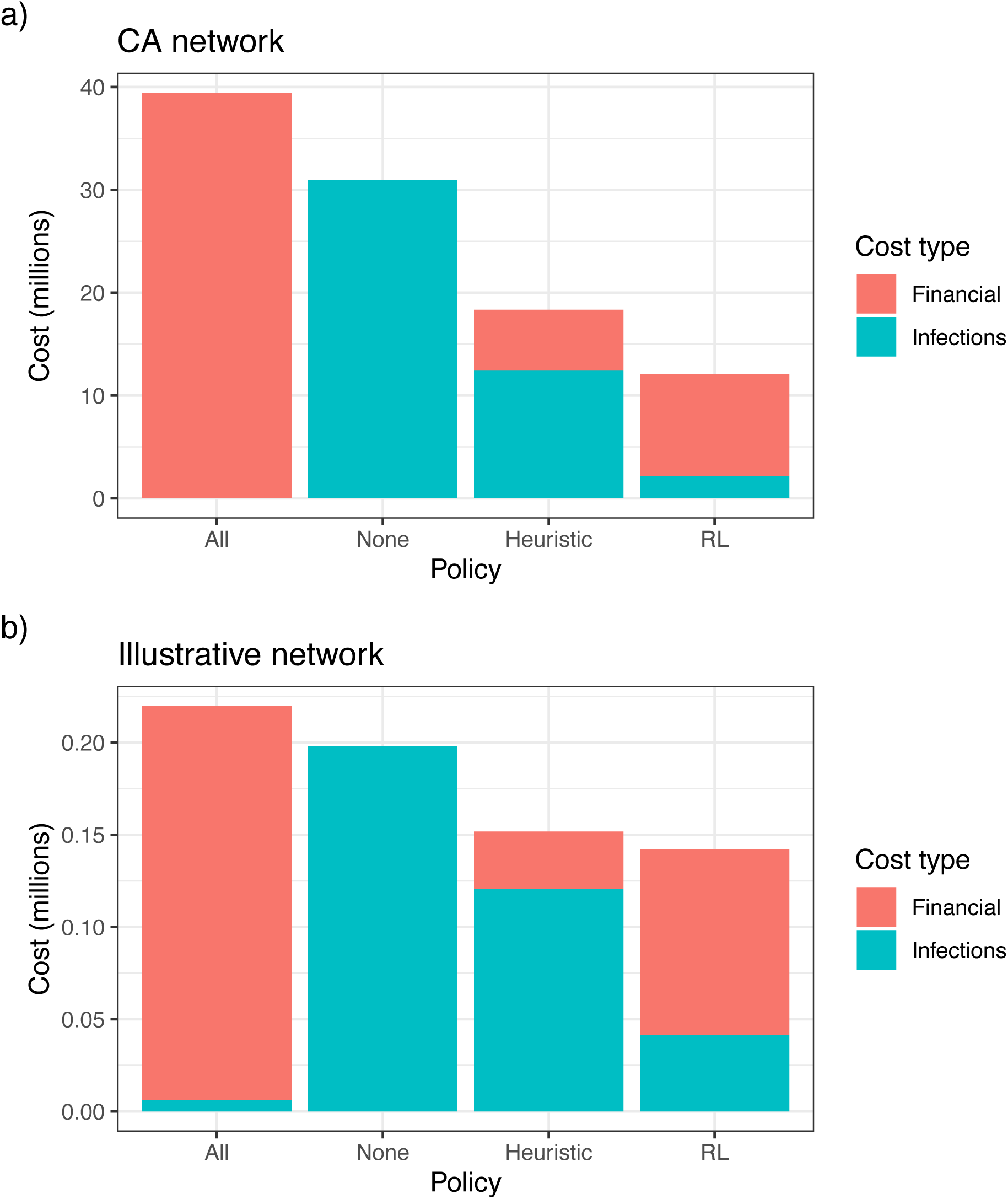
Performance of control strategies. We simulated outcomes including infections and control measure costs associated with each of four control strategies 1,000 times. These strategies included all communities and prisons implementing control measures at maximum for the model duration, none of them implementing control measures for the model duration, a heuristic strategy, and an RL strategy. We converted both infections and control measure costs to units of infections using a WTP threshold and plotted their sum averaged over the 1,000 replications.

To interpret the RL strategy for the California network, we fit an XGBoost model to predict the average expenditure for each community and an XGBoost model to predict the average expenditure for each prison while following this strategy. From these XGBoost models, we generated variable importance and partial dependence plots (Figure 3, Figure 4). From the variable importance plots, we found the most important variables for predicting average expenditure were different for communities versus prisons. From the partial dependence plots, we observed the marginal effects of each variable on average expenditure. For communities, the RL policy’s resource allocation focused on those with higher betas, and, to a much lesser extent, those whose members more frequently visited other communities and those with fewer people. Beta was likely particularly important for communities as communities with higher betas are more likely to have severe outbreaks, which can precipitate outbreaks in other communities and prisons. For prisons, the RL policy’s resource allocation focused on those with more staff per incarcerated person, and, to a lesser extent, those with more incarcerated people and those with more turnovers (i.e., incarcerations and releases) per incarcerated person. Least importantly, it allocated more resources to prisons with higher betas up to some point at which slightly fewer resources were allocated. Staff per incarcerated person and number of incarcerated people were likely particularly important for prisons as prisons with higher values of these variable are more likely to experience and then spread outbreaks. Beta was likely much less influential for prisons as all the prisons had relatively high betas.

**Figure 3.**
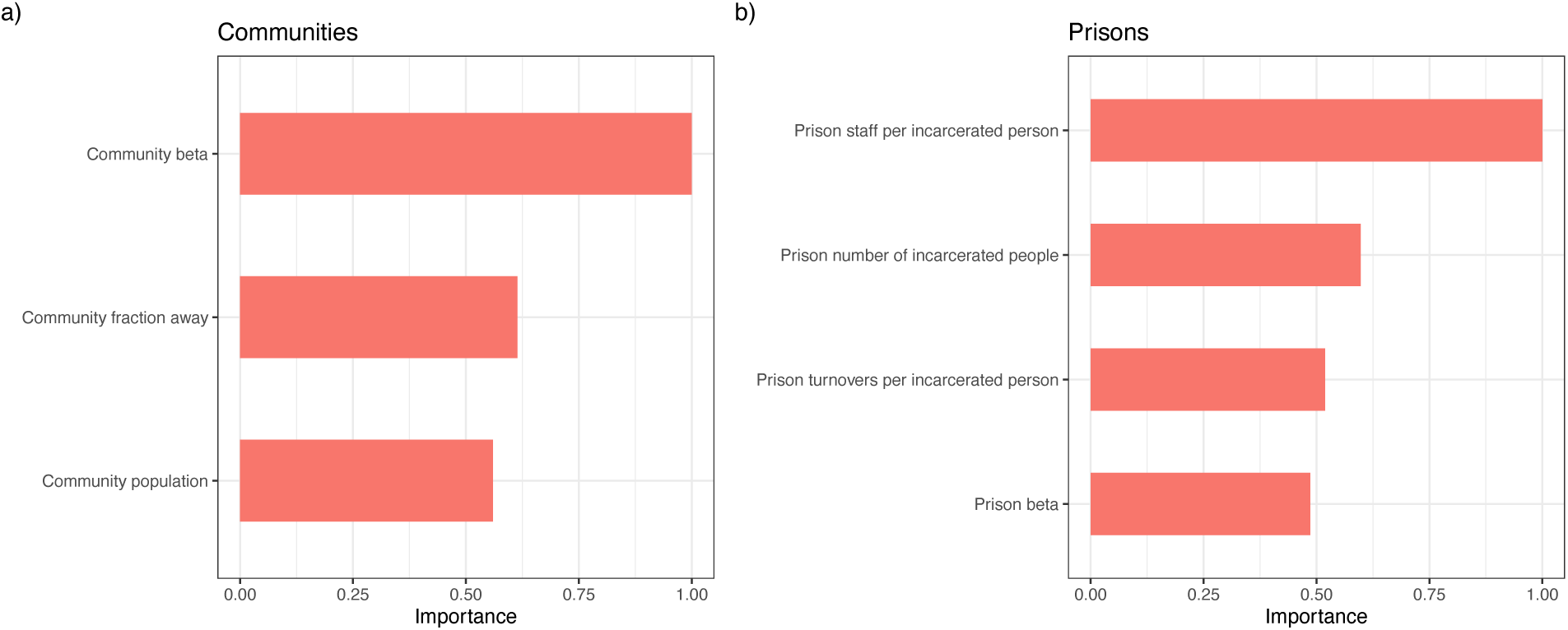
Variable importance plots. To interpret the RL strategy for the California network, we calculated the average expenditure on control measures per person per week per control measure for each community and prison. We then fit XGBoost models for communities and prisons, respectively, to predict average expenditure as a function of community and prison variables. We plotted variable importance.

**Figure 4.**
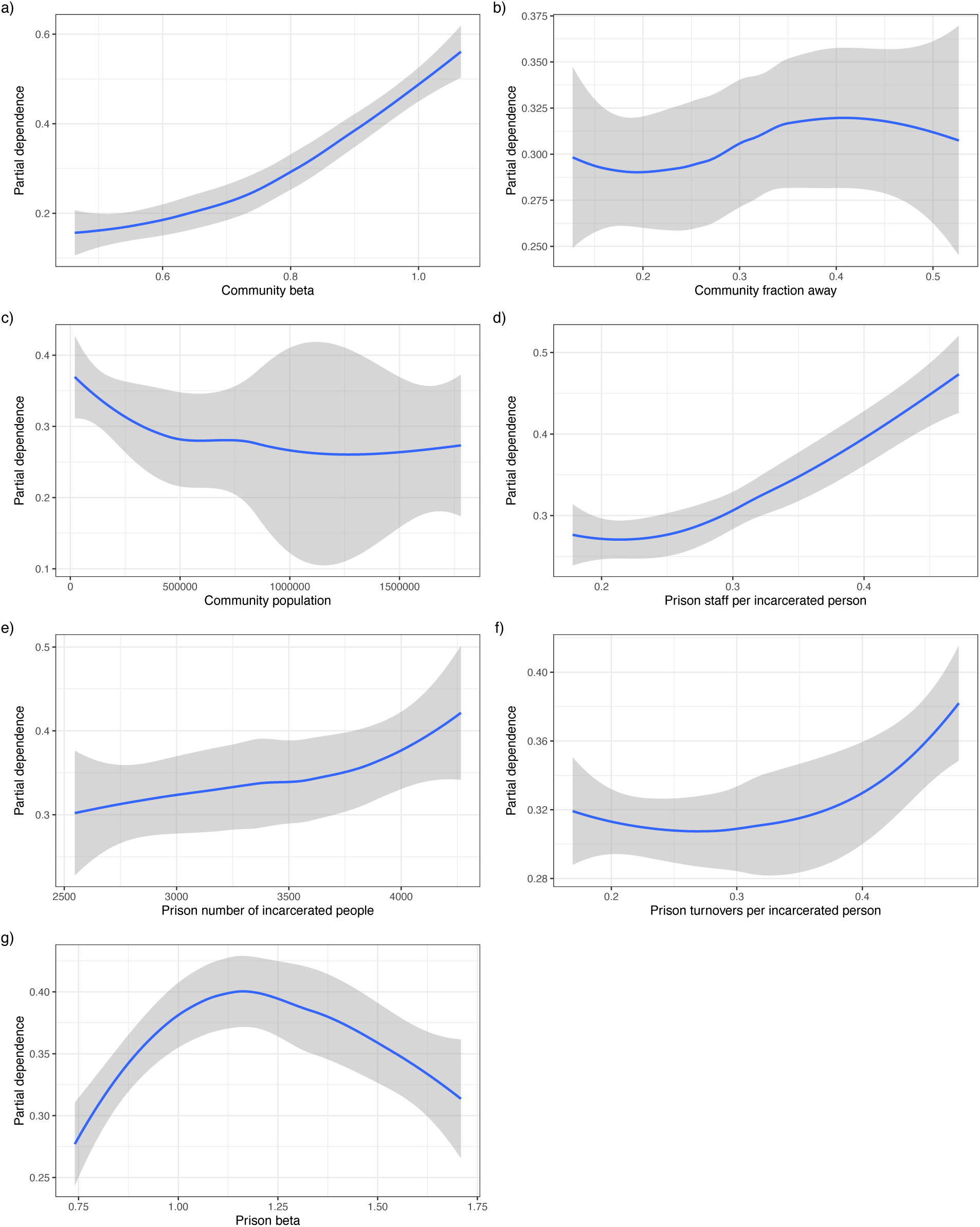
Partial dependence plots. We generated partial dependence plots for the models described in Figure 3. Plots a-c show the results for the community model while plots d-g show the results for the prison model. The y-axes were scaled by a factor to assist with readability.

The superiority of the RL strategy was robust to changes in following the recommended actions exactly and changes in disease parameters for the California network (Figure 5a). For the base case, varying the fidelity of implementing a strategy’s recommended actions, and varying disease parameters, the RL strategy was best in 99.7%, 99.5%, and 98.9% of the 1,000 replications, respectively.

**Figure 5.**
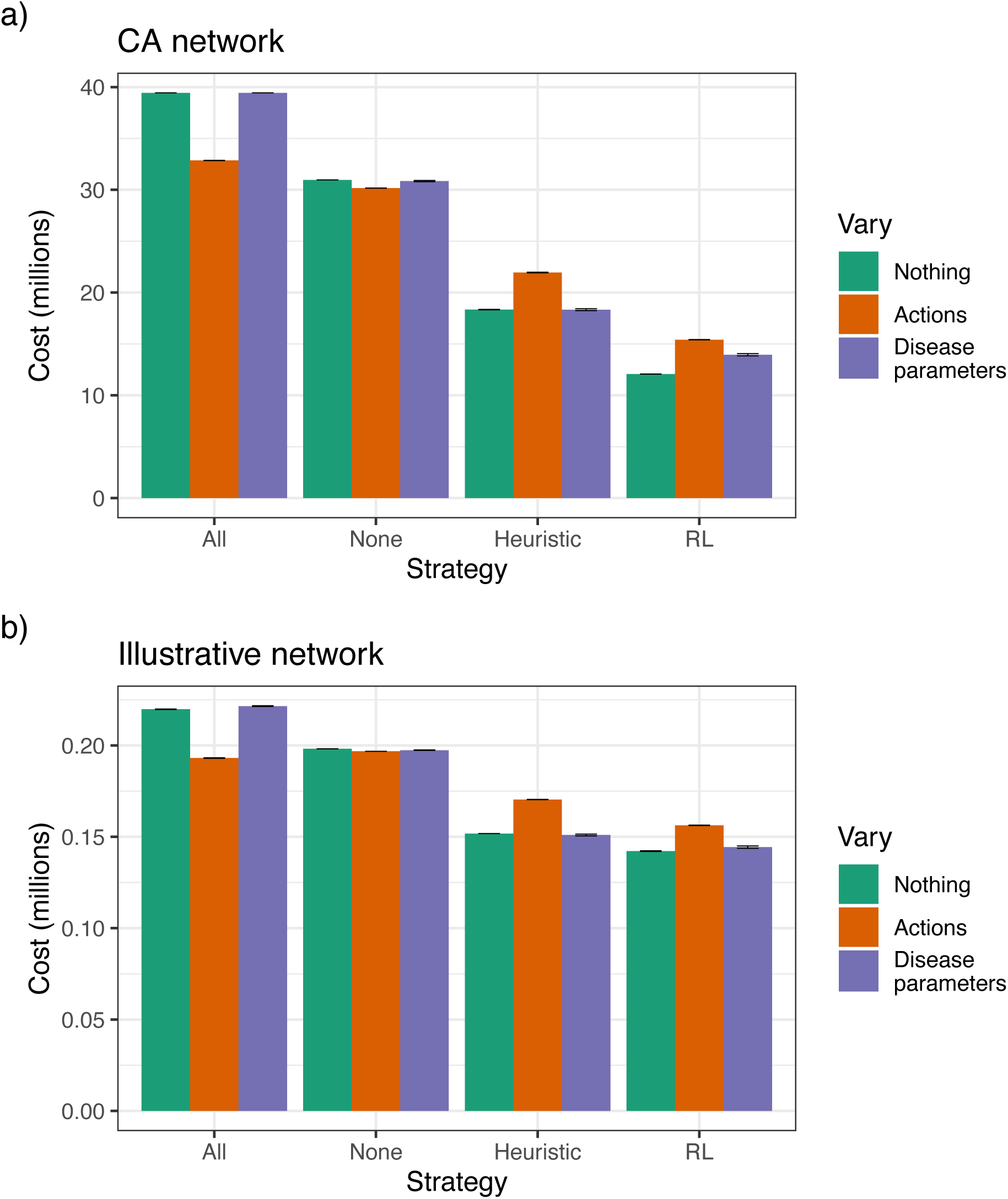
Sensitivity analysis. We repeated the analysis from Figure 2 for three cases. First, nothing was varied such that the height of each bar was equal to infections plus financial costs from Figure 2. Second, we randomly varied each action recommended by each control strategy such that there were equal chances of plus one level (+20%), no change, and minus one level (- 20%). Finally, we randomly varied disease parameters in each iteration as described in the methods section. The error bars show plus and minus the standard error.

### Illustrative network

The California network was large and complex. To provide additional credibility to our findings and allow for easier interpretation of strategies, we also considered a smaller, illustrative network. The RL strategy offered better performance than the other control strategies for the illustrative network (Figure 2b). We projected that following control all, control none, heuristic, and RL strategies would result in costs of 0.22, 0.20, 0.15, and 0.14 million infections, respectively. Again, these costs are the sum of the actual infections and the financial costs of the control measures, which were first converted to units of infections using a WTP threshold as described in the Methods section.

To interpret the RL strategy for the illustrative network, we visualized the epidemic curves and the RL policy’s recommended actions per community and prison over time (Figure S2). These plots suggest that the reason the RL strategy is able to perform better is because it exploits the network structure (Figure 1b) and is able to coordinate actions of each community and prison. In terms of exploiting the network structure, first, we found the agent allocated the most resources to the middle prison to disrupt flow of the epidemic across the entire network, as the middle prison bridged the communities on the left in which the epidemic started with those on the right. Second, when the outbreak reached the middle prison from its start in the leftmost community, the agent substantially increased testing in the prison on the right and NPI usage in the communities on the right. Finally, the agent used substantial NPIs and testing in the middle prison before an outbreak occurred at that prison. In terms of coordination, our findings suggest the agent is trying to mitigate prison amplification dynamics. For example, it starts mitigation in the middle prison before the outbreak occurs at that prison, and, once it does occur, it initiates control measures in the communities and prison to the right of the middle prison before they have outbreaks.

The superiority of the RL strategy was robust to changes in following the recommended actions exactly and changes in disease parameters for the illustrative network (Figure 5b). For the base case, varying the fidelity of implementing a strategy’s recommended actions, and varying disease parameters, the RL strategy was best in 100.0%, 100.0%, and 90.9% of the 1,000 replications, respectively.

## Discussion

We developed a simulation model of an epidemic spreading across a network of communities and correctional facilities. We parameterized this model for COVID-19 spread on 1) California communities and prisons and 2) an illustrative network of communities and prisons.

We then compared different control strategies on these networks, and we found that in both cases the RL strategy substantially outperformed the other strategies. For the RL strategy, we identified differences in the factors governing allocation of resources to communities versus prisons, and we found evidence of geo-temporal patterns consistent with trying to mitigate prison amplification dynamics. Finally, we found the superiority of the RL strategy was robust to changes in following the recommended actions exactly and changes in disease parameters.

Our work highlights how control of an epidemic on a network of communities and correctional facilities can be robustly improved through the use of modern quantitative methods, such as RL.^18,19^ Policy makers should consider investing in the further development of such methods and using them for future epidemics. Beyond that, we offer qualitative insight into different factors that might inform resource allocation to communities versus prisons during future epidemics.

Our analysis has several limitations. First, due to our use of a stochastic compartmental model, we assumed homogeneous mixing of people within each community and prison. While this is an approximation of actual mixing dynamics, it is a common assumption when modeling respiratory infectious diseases in both communities and correctional facilities. Second, we assume illustrative costs and a willingness-to-pay threshold per infection averted. Further work should characterize these values for specific applications before such a framework is used in practice. Third, we only considered communities and prisons due to time constraints and data availability. However, jails and other high density institutional settings could easily be added to the network in our framework.

To conclude, for both the California and illustrative networks, the RL strategy to control epidemic spread on networks of communities and correctional facilities robustly offered the best performance. It substantially outperformed the heuristic strategy, which is similar to strategies that were typically used during the COVID-19 epidemic. We also derived qualitative insights into the behavior of the RL strategy. Taken together, our work offers tools and insights that can help guide resource allocation to communities and correctional facilities during future epidemics.

## Ethical considerations

Stanford University’s Institutional Review Board approved the use and analysis of primary California Department of Corrections and Rehabilitation (CDCR) data (IRB-55835).

## Data Availability

Input data and code for replication and extension of our analysis will be available via GitHub concurrent with publication.

