## Supplement for "Reinforcement learning-based control of epidemics on networks of communities and correctional facilities"

This supplement provides additional details on model inputs, methods, and results.

**Text S1. Detailed description of simulation model.**

We developed a stochastic, metapopulation SEIDR model of infections occurring within and between communities and correctional facilities. For ease of terminology, we assumed the correctional facilities were prisons in this section. The model is a metapopulation model in that we modeled a network of prisons and communities, which each have their own underlying population of people. These underlying populations were composed of incarcerated people (by prison), staff (by prison and community), and free-living people (by community). The model is an SEIDR model in that the simulation tracked the movement of people in these underlying populations between the mutually exclusive and collectively exhaustive health states: susceptible (S), exposed (E), infectious (I), detected (D), and recovered (R). At the outset, all people were susceptible except for a small number of infectious individuals in one community forming the seed of the outbreak. The model was stochastic in that we simulated transitions between the health states using the tau-leap method, which is an approximate method for stochastic simulation based on Gillespie’s algorithm, with a time step of a tenth of a day.^1^

People in the communities and prisons are interconnected and can interact in several ways. First, we modeled staff spending part of their time in the prison in which they worked and part of their time as members of their home community. Second, we modeled incarcerations and releases to/from prisons. Third, we modeled community mobility (e.g., daily commutes between communities). The details of these interactions and data sources are described subsequently.

We modeled transitions between health states for incarcerated people (by prison), staff (by prison and community), and free-living people (by community). For these transitions, susceptible people become exposed (infected but not yet infectious) based on betas for each community (β_C,i_) and prison (β_P,i_) assuming homogeneous mixing of people within each community and prison. Exposed people become infectious at a rate σ. Infectious people become detected based on detection rates for each community (d_C,i_) and prison (d_P,i_). Detected people are then isolated, reducing their ability to transmit based on a reduction factor d_red_. Infectious and detected people become recovered at a recovery rate (γ).

In each time step, we first computed the S, E, I, D, and R states for the prison and community populations based on these states for incarcerated people, free-living people, and staff. The prison populations consisted of the incarcerated people plus the number of staff for those prisons weighted by the fraction of time the staff spend at the prisons. The community populations consisted of the free-living populations weighted by where those people spend time plus the number of staff for those communities weighted by the fraction of time the staff spend at those communities. C_mob,i,j_ gives the fraction of time people from community i spend in community j. Staff spend a fraction of time in the free-living population versus in the prison where they work (f_time_com_).

We calculated the forces of infection for the prisons, communities, incarcerated people, free-living people, and staff according to the following equations:

$$\lambda_{P,i}=\beta_{P,i}{(I}_{P,i}+{d_{red}D}_{P,i})/N_{P,i}$$

$$\lambda_{C,i}=\beta_{C,i}{(I}_{C,i}+{d_{red}D}_{C,i})/N_{C,i}$$

$$\lambda_{incarcerated,i}=\lambda_{P,i}$$

$$\lambda_{free,i}=\sum_{k=i}^{N} C_{mob,i,k}\lambda_{C,k}$$

$$\lambda_{staff,i,j}=f_{time\_com}\sum_{k=i}^{N} C_{mob,i,k}\lambda_{C,k}+{{(1-f}_{time\_com})\lambda}_{P,j}$$

In these equations, “I” denotes the number of undetected infectious people, “D” denotes the number of detected infectious people, “N” denotes the number of people, “P” denotes prison, “C” denotes community, “i” denotes the community or prison number (arbitrarily assigned index), “k” and “j” denote community or prison numbers when other indices are needed, and “f” denotes fraction. The “N” variables reflect the number of people at each community or prison at any given time.

We calculated the number of new exposed people for incarcerated people, free-living people, and staff based on the lambdas and susceptible populations. We calculated the number of new infectious people based on the rate σ and exposed populations. We calculated the number of new detections based on the rate d and the infectious but undetected populations. We calculated the number of new recoveries based on the rate γ and the infectious populations including both detected and undetected people. We updated the S, E, I, D, and R states for these populations.

We assumed that incarcerations and releases between each community and prison were equal such that the number of people in each community and prison was constant. We assumed that incarcerations and releases occur once per week. To model them, for each week, we first sampled the number of incarcerations and releases between each prison-community pair by sampling from Poisson distributions with lambdas equal to the mean weekly numbers. For each prison-community pair, we then sampled the SEIR states of those entering and leaving the prison by sampling from multivariate hypergeometric distributions parameterized by the number of people in each state in the community and prison, respectively (computed from the model at each time). We omitted people in the D state from these distributions as we assumed detected infectious people would not be incarcerated or released while isolating.

For control measures, we considered non-pharmaceutical interventions (NPIs) and testing. We assumed each of these strategies had a maximum level of use (NPI_max_ and test_max_). NPI_max_ gives the maximum reduction in beta (e.g., a value of 40% means that the beta is reduced 40% at max). This reduction in beta might occur via a reduced number of contacts and/or probability of transmission given contact. test_max_ gives the maximum testing rate. We allowed for six different levels of use for each control measure between no use and maximum use (0%, 20%, 40%, 60%, 80%, and 100%). For testing, we assumed some free, low-level of testing was always available to avoid the problem where no testing suggests no infections. We allowed control strategies (either heuristic or RL) to update their use of control measures for each community and each prison weekly.

To provide additional clarification on the control measures, if NPI_max_ was equal to 40% and the level of use in a prison was 60%, the multiplier on beta for that prison would be 1 - 0.4*0.6 = 0.76. This reduced beta would lower the force of infection for that prison, which would lower the forces of infection for incarcerated people and staff at that prison.

**Text S2. Epidemiologic parameters.**

We selected epidemiologic parameters based on published literature to reflect those for California during the early phases of the COVID-19 epidemic. See Table S1. The community-specific and prison-specific betas were calculated based on R0s estimated from empirical time series data. For communities, we used empirical time series data on infections from covidestim.^2^ We calculated Rt values for each community each day in the first month of available data (early 2020) and computed their means to estimate R0s. For prisons, we used empirical time series data based on cases from CDCR.^3^ We identified the first outbreak as the date with more than 5 cases. We then calculated Rts from the day with the first case in the period one week before this date to one week after this date and computed their means to estimate R0s. Since there were a few outliers, we applied a rule such that if any community had an R0 greater than the median prison R0, it was set to the median prison R0. And, if any prison had an R0 less than the median community R0, it was set to the median community R0. We calculated Rts for each community and prison using the R0 package in R and the Wallinga and Teunis method^4^ with correction for estimation in real time^5^ (Figure S3). Betas were calculated from the R0s per standard practice.

**Text S3. Connectivity parameters.**

We assumed incarcerated people spend all their time in the correctional facility in which they are incarcerated. We obtained data on the number of people incarcerated in each California prison from CDCR as of early 2020.^3^ We obtained data on weekly releases from each prison to each county from CDCR for 2020.^3^

For simplicity, we assumed staff spend 50% of their exposure time in the correctional facility in which they work and 50% as a member of their home community. We define exposure time as the effective time during which transmission to and from these people could occur. We obtained data on the number of staff in each California prison and the communities in which they reside from CDCR as of early 2020.^3^

We determined community mobility based on data from SafeGraph, which tracks movement of millions of people with mobile devices in the US.^6^ SafeGraph defines people by their home community (where they most frequently spend the night) and tracks the movement of these people to places within and outside of their home community. Our estimates were based on the weekly census block group (CBG) to point of interest (POI) visitor flow dataset. CBGs are geographical areas that are finer than zip codes. POIs represent physical locations that may be of interest and include a very wide array of places (e.g., restaurants, retail stores, transit stops, ATMs, etc.). Specifically, we used a dataset that had aggregated this information to weekly county-to-county visitor flows and corrected for the number of cell phone users in the dataset such that the visitor flows were for all people in CA as opposed to just those with cell phone data.^7^ To be clear, visits could be made to a person’s county of origin, which were the most common in the dataset. We used data from the first week of April 2019 to represent the mobility at the start of the COVID-19 epidemic. To compute the community mobility matrix used in our model, we normalized the visitor flows from each county such that they summed to one (Figure S4).

**Text S4. Costs parameters.**

We selected cost parameters for illustrative purposes (Table S2). There are several points worth noting about our choices. First, we set the costs parameters such that the cost of using all control measures at maximum intensity (defined by NPI_max_ and test_max_ as described above) for the full model duration was approximately equal to 1.25 times the cost of the infections that would occur if no control measures were used. We made this choice as a substantial imbalance results in a trivial optimal policy of using no or maximum mitigation in all communities and prisons. Second, we set the cost parameters such that the cost of using the two control measures at the same intensity was equal. We made this choice as a substantial imbalance results in a trivial optimal policy of using one control measure but not the other. Finally, we assumed the cost of the control measures was non-linear in the intensity of their use. We made this choice as it becomes ever more costly to achieve higher intensities of control measures as additional supplies are needed and/or people less likely to comply need to be convinced.

**Text S5. Estimation of infections and cumulative infections.**

We estimated weekly infections, including both detected and undetected infections, for each community and prison. We estimated this by accounting for the fact that people are undetected if they recover before moving to the detected state. Recoveries occur at rate γ while detections occur at rate d, which are both known. Thus, if D detections occur in one week, we can approximate the total number of infections that would have occurred based on these D detections as D * (γ + d) / d. We estimated cumulative infections that have occurred in community or prison i by summing this quantity over time up to the current week T:

${cum\_inf}_{i}=\sum_{t=1}^{T} D_{i,t}(\gamma+d_{i,t})/d_{i,t}$.

**Text S6. Heuristic strategy search.**

We evaluated a heuristic control strategy in which each community or prison implements control measures at a certain intensity based on the per-capita incidence of detected infections in that community or prison and a community or prison-specific threshold. To be clear, there was one threshold and one intensity for all communities, and there was one threshold and one intensity for all prisons.

We determined the thresholds and intensities of control measures for communities and prisons, respectively, through exhaustive search. For both communities and prisons, the thresholds for estimated incidence of infections considered were 0%, 0.5%, 1%, 2.5%, 5%, and 10%. The control measure intensities considered were 0%, 20%, 40%, 60%, 80%, and 100% to mirror those considered during RL. Thus, we tried 6*6*6*6 = 1,296 possible combinations of parameter values. We projected outcomes for each combination of parameter values 5 times and computed the mean reward. We computed the reward as described in the “Reinforcement learning method” section of the main paper. We selected the optimal heuristic strategy as the one that gave the highest reward (i.e., minimized infections and cost of the control measures). We repeated the heuristic strategy search a second time to ensure that the same heuristic strategy was optimal.

**Text S7. Hyperparameter tuning.**

We tuned hyperparameters using Optuna.^8^ The possible hyperparameter values considered in the search are shown in Table S3. We used a sampler using the Tree-structured Parzen Estimator (TPE) algorithm, a median pruner, a budget of 50 trials, a maximum of 104,000 steps, and 20 evaluation episodes.

**Table S1.** **Epidemiologic parameters.**

| **Parameter** | **Value** | **Source** |
| --- | --- | --- |
| *CA network* |  |  |
| Prison betas | Prison specific | Calculated from CDCR^3^ |
| Community betas | Community specific | Calculated from covidestim^2^ |
| *Illustrative network* |  |  |
| Prison betas | 1.92 | Based on those for CA network but slightly elevated since no heterogeneity |
| Community betas | 0.96 | Based on those for CA network but slightly elevated since no heterogeneity |
| *Shared parameters* |  |  |
| Rate exposed to infectious | 1/3 per day | Goldhaber-Fiebert *et al.*^9^ |
| Recovery rate | 1/3.12 per day | Goldhaber-Fiebert *et al.*^9^ |
| Infections seeded | 100 | Assumed |
| Reduction in infectiousness upon detection due to isolation | 90% | Assumed |
| Maximum NPI level (percent reduction in β) | 50% | Assumed |
| Maximum testing rate | 1/3.5 per day | Assumed |

**Table S2.** **Cost of mitigation strategies per person per week for each level.** The numbers here are multiplied by 52*2 for ease of comparison. The 52 comes from the model duration of 52 weeks, and the 2 comes from the 2 control measures. For example, the cost of having both mitigation strategies at 80% for one person-week would be (0.5+0.5)/(52*2).

| **Mitigation level** | **Cost (cost units)** |
| --- | --- |
| 0% | 0.000 |
| 20% | 0.025 |
| 40% | 0.050 |
| 60% | 0.150 |
| 80% | 0.500 |
| 100% | 1.000 |

**Table S3.** **Hyperparameter values considered for RL.** Parameter values used in the search were based on those recommended by Stable Baselines3.^10^ Some parameters are multiples of 52 since that is the number of steps per episode.

| **Parameter** | **Parameter type** | **Possible values** |
| --- | --- | --- |
| batch_size | categorical | [52*16, 52*32, 52*64] |
| n_steps | categorical | [52*16, 52*32, 52*64, 52*128, 52*256] |
| gamma | categorical | [0.9, 0.95, 0.98, 0.99, 0.995, 0.999, 0.9999] |
| learning_rate | float | min=1e-6, max=1e-3, log=True |
| ent_coef | float | min=0.00000001, max=0.1, log=True |
| clip_range | categorical | [0.1, 0.2, 0.3, 0.4] |
| n_epochs | categorical | [1, 5, 10, 20] |
| gae_lambda | categorical | [0.8, 0.9, 0.92, 0.95, 0.98, 0.99, 1.0] |
| max_grad_norm | categorical | [0.3, 0.5, 0.6, 0.7, 0.8, 0.9, 1, 2, 5] |
| vf_coef | float | min=0, max=1 |
| net_arch_type | categorical | [dict(pi=[256, 256], vf=[256, 256]),  dict(pi=[512, 512], vf=[512, 512]),  dict(pi=[1024, 1024], vf=[1024, 1024])] |
| activation_fn_name | categorical | ["tanh", "relu"] |

**Table S4. Hyperparameter values considered for models to predict average expenditure on control measures for each community and prison.** We used the caret package in R to perform hyperparameter tuning using leave-one-out cross validation. Parameter values used in the search were based on those recommended by caret.^11^

| **Parameter** | **Possible values** |
| --- | --- |
| *XGBoost* |  |
| eta | 0.3, 0.4 |
| max_depth | 1, 2, 3 |
| colsample_bytree | 0.6, 0.8 |
| subsample | 0.5, 0.75, 1.0 |
| nrounds | 50, 100, 150 |
| *Random forest* |  |
| splitrule | variance, extratrees |
| mtry | community: 2, 3  prison: 2, 3, 4 |
| *Glmnet* |  |
| alpha | 0.10, 0.55, 1.00 |
| lambda | community: 0.02, 0.002, 0.0002  prison: 7.5*10^-3^, 7.5*10^-4^, 7.5*10^-5^ |

**Table S5. Performance of models to predict average expenditure on control measures for each community and prison.** For reference, the standard deviation of the community and prison targets were 0.16 and 0.22, respectively. Abbreviations: RMSE, root mean square error; MAE, mean absolute error.

| **Model type** | **RMSE** | **MAE** |
| --- | --- | --- |
| *Community* |  |  |
| XGBoost | 0.06 | 0.04 |
| Random forest | 0.07 | 0.05 |
| Glmnet | 0.11 | 0.09 |
| *Prison* |  |  |
| XGBoost | 0.11 | 0.09 |
| Random forest | 0.13 | 0.10 |
| Glmnet | 0.21 | 0.16 |

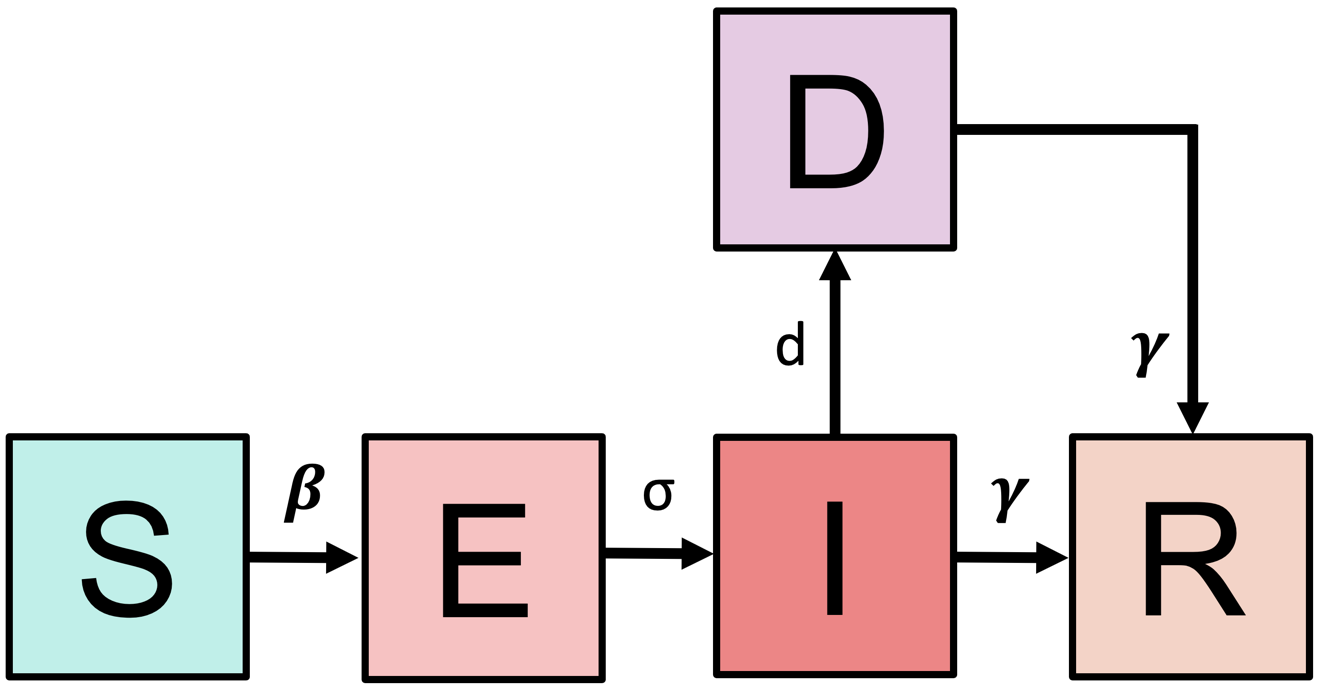
**Figure S1. Model schematic.** The model tracks susceptible-exposed-infectious-detected-recovered (SEIDR) states for incarcerated people in each prison, correctional staff by the prison in which they work and their home community, and free-living people in each community. These models are not isolated, and people interact with each other based on the network connections as described in Text S1. If the models were isolated, susceptible people would become exposed based a rate β. Exposed people would become infectious based on a rate σ. Infectious people would be detected based on a rate d. Infectious and detected people would recover based on a rate γ.

**Figure S2. Interpretation of RL policy for illustrative network.** We visualized the a) percent of people infected at each community and prison, and the b) RL policy’s recommended actions over time for one simulation. Other simulations were slightly different but showed the same qualitative patterns. For plot a, Inf0_p denotes infections for prison 0 while Inf0_c denotes infections for community zero. For plot b, B0_p denotes NPIs usage for prison 0 (B because NPIs effects betas) while d0_p denotes testing usage for prison 0 (d because testing effects the detection rate).

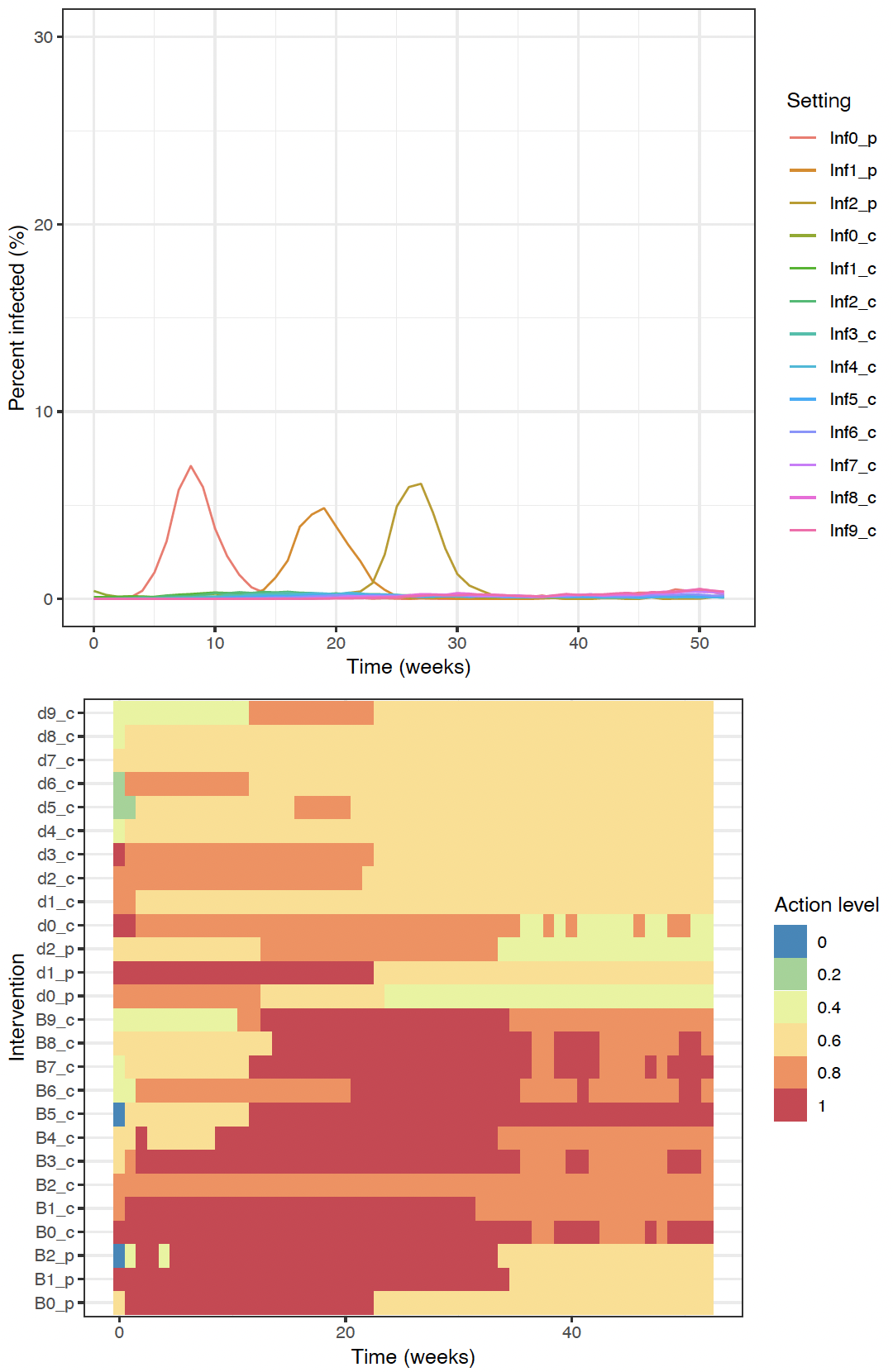

a)

b)

**Figure S3. Estimated R0s.** We plotted our estimated R0s for prisons and communities, respectively.

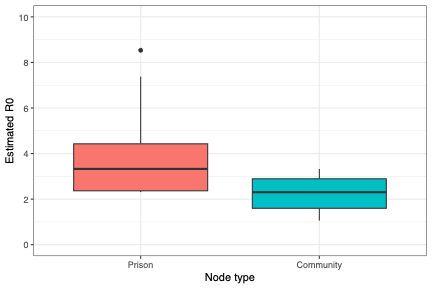

**Figure S4. Estimated community connection strengths.** We plotted the connection strengths between each community and each other community.

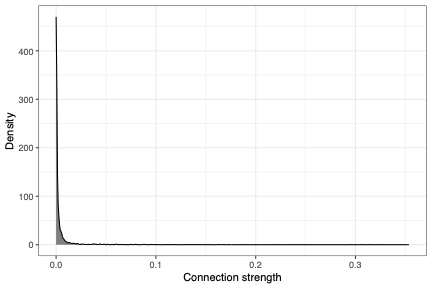
